# Evaluation of a throat spray with lactobacilli in COVID-19 outpatients in a randomized, double-blind, placebo-controlled trial for symptom and viral load reduction

**DOI:** 10.1101/2022.03.17.22272401

**Authors:** Ilke De Boeck, Eline Cauwenberghs, Irina Spacova, Thies Gehrmann, Tom Eilers, Lize Delanghe, Stijn Wittouck, Peter A. Bron, Tim Henkens, Imane Gamgami, Alix Simons, Ingmar Claes, Joachim Mariën, Kevin K. Ariën, Diana Bakokimi, Katherine Loens, Kevin Jacobs, Margareta Ieven, Patricia Bruijning-Verhagen, Peter Delputte, Samuel Coenen, Veronique Verhoeven, Sarah Lebeer

## Abstract

**Objectives:** Primary care urgently needs treatments for COVID-19 patients because current options are limited, while these patients account for more than 90% of the people infected with SARS-CoV-2.

**Methods:** We evaluated a throat spray containing three *Lactobacillaceae* strains with broad antiviral properties in a randomized double-blind placebo-controlled trial. Seventy-eight eligible COVID-19 patients were randomized to verum (n=41) and placebo (n=37) within 96 hours of positive PCR-based SARS-CoV-2 diagnosis and per-protocol analysis was performed. Symptoms and severity were reported daily via an online diary. Combined nose-throat swabs and dried blood spots were collected at regular time points in the study.

**Results:** The daily reported symptoms were highly variable, with no added benefit for symptom resolution in the verum group. Specific monitoring of the applied lactobacilli strains showed that they were detectable via microbiome (27%) and qPCR analysis (82%) of the verum group. Their relative abundances were also negatively correlated with the acute symptom score. At the end of the trial, a trend towards lower SARS-CoV-2 viral loads was observed for the verum group (2/30, 6.7% positive) compared to the placebo group (7/27, 26% positive) (p = 0.07).

**Conclusions:** Despite a trend towards lower SARS-CoV-2 viral loads at the end of the trial and a negative correlation between relative abundances of the applied lactobacilli in the microbiome and acute symptoms, we did not observe a significant effect on overall symptom score for the verum group. This suggests that studies with earlier application of the spray in larger study populations are needed to further assess application potential.

## Introduction

During the COVID-19 pandemic, most research and clinical trials on treatment options have been conducted in hospitalized patients. This especially applies to intervention studies, which are routinely executed in a hospital setting in critically ill patients. However, only 10-20% of COVID-19 patients need medical care in hospitals [1]. While these numbers vary depending on the dominating SARS-CoV-2 variant, this means that the majority of COVID-patients have mild to moderate symptoms, are not hospitalized, and depend only on treatments such as antiflogistics and analgesics [1,2]. Nevertheless, these milder cases exert a significant burden on healthcare professionals in primary care [2,3]. In addition, asymptomatic and presymptomatic transmission is the main driver for to others [4]. Respiratory viral infections can have severe health consequences due to imbalanced immune activation and bacterial co-infections associated with airway tissue disruption and severe inflammation [5]. This clearly shows the urge for more treatment and/or prevention options in COVID-19 outpatients, which can improve different aspects of the disease: symptom relieve, transmission reduction and decreased hospitalization.

Microbiome or probiotic therapy is an emerging alternative treatment option for respiratory viral diseases based on the potential multifactorial action of beneficial bacteria in the airways [6]. While oral administration of such microbiome therapeutics or probiotics remains most common [7], this route relies on systemic effects to ameliorate respiratory infections. Also during the current COVID-19 pandemic, oral administrations targeting the gut have already been explored [8–10]. Alternatively, topical application of rationally-selected probiotics in the airways might offer several advantages [11], as this could lead to direct blocking or inhibition of respiratory viruses [12], and direct immune modulation at the site of infection and inflammation [13,14]. Indeed, the probiotic definition is not limited to the gut [15]. We recently developed a microbiome-modulating throat spray with three *Lactobacillaceae* strains that were selected based on their safety and *in vitro* multifactorial modes of action on the key aspects of viral infection and disease, and their ability to thrive in the human respiratory tract of healthy volunteers [16]. Yet, microbiome therapy with live bacteria has several challenges, such as formulation and selection of target patient population.

Here, we evaluated the clinical potential of this throat spray with *Lacticaseibacillus casei* AMBR2, *Lacticaseibacillus rhamnosus* GG, and *Lactiplantibacillus plantarum* WCFS1 against COVID-19 in a randomized, double-blind, placebo-controlled trial in COVID-19 outpatients exhibiting mild-to-moderate symptoms. Specifically, we monitored impact on symptom severity, time to improvement, viral load, anti-SARS-CoV-2 antibodies and the respiratory microbiome in an out-of-hospital setting. This trial relied on self-sampling and included collection of combined nose-throat swabs, fingerprick blood samples and reporting of symptom and severity via an online diary.

## Methods

### Clinical trial design

A double-blind, placebo-controlled clinical trial was performed with a microbiome throat spray in COVID-19 outpatients within 96 hours after a positive PCR test in government facilities (see more details in supplementary methods). Approval was obtained from the committee of medical Ethics (UZA/UAntwerpen, B3002021000018 and NCT04793997). Informed consent was obtained from all participants prior to inclusion. Verum and placebo sprays were supplied by Yun NV (Niel, Belgium) (supplementary methods). Randomization occurred in blocks of six patients with stratification for age and gender and was done by the responsible clinicians (see supplementary methods for more details).

### Study procedures

Patients were asked to use the verum spray or placebo for 14 days, with one week of follow-up, and filled in an online diary via Qualtrics (Qualtrics, Provo, UT, USA) (detailed description in Figure S1). Ten common COVID-19 symptoms were monitored, according to [17]. Different symptom summary scores were compared between the verum and placebo groups, and time to improvement was evaluated based on the timepoint when participants reached their symptomatic tipping point (supplementary methods). Combined nose/throat swabs for microbiome analysis, determination of SARS-CoV-2 viral loads and detection of administered *Lactobacillaceae* strains via qPCR and MiSeq amplicon sequencing were self-sampled, as well as blood fingerprick samples (dried blood spots) to analyze SARS-CoV-2 IgG antibodies (Figure S1 and supplementary methods).

### Outcomes

The primary clinical outcome of this trial was the change in severity of COVID-19 infection symptoms after using the microbiome spray. The secondary study outcomes included: (i) Change in duration (time to improvement) of COVID-19 infection symptoms after using microbiome spray, (ii) Change in absolute level of SARS-CoV-2 particles after using microbiome spray, (iii) Change in absolute numbers of specific bacterial pathogens after using microbiome spray, and (iv) Change in microbiome of nose/throat region after using microbiome spray.

Finally, some explorative (post-hoc) analyses were included: (i) relation of viral load with reported symptoms, (ii) colonization of the administered strains in the airways and (iii) correlation of the microbiome with several variables.

### Statistical analyses

Per protocol analysis was performed on participants that completed the study and provided samples on all timepoints. Standardized scores (z scores) were used for the analysis of the primary outcome. The distributions of the severity score for four different symptom summary scores (see supplementary methods) on every day between both treatment groups were compared. Kaplan meier survival analysis was performed using the R survival package. The symptomatic tipping point was taken as the event, and the time until occurrence was tested in the different treatment groups. Differences between symptom scores in covid-positive and -negative participants (based on PCR) were tested using a random effect model, symptom ∼ covid + (1|participant). P-values were adjusted for multiple testing using Bonferroni. Differential bacterial abundances between treatment groups were tested with a random effect model with CLR(ra) ∼ treatment + plate + qubit_score + library_size + (1|participant), where CLR(ra) is the centered-log-ratio transformed relative abundance of a given bacteria, and plate, qubit_score and library size constitute technical confounders. Effect sizes for the treatment group were calculated for timepoints T2 and T3, at which the participants were using the spray.

## Results

### Set-up of a placebo-controlled intervention trial in mild-to-moderate COVID-19 patients and assessment of study compliance and self-sampling

The trial was conducted from February 24, 2021 to April 30, 2021 at the University of Antwerp. Seventy-eight eligible patients were randomized, of which 41 were allocated to the verum spray and 37 to the placebo spray. Figure 1 depicts patient recruitment and enrollment. Fourteen participants dropped out during the trial (7 verum, 7 placebo; reasons in Figure 1) and per protocol analysis of the remaining participants that provided samples on all timepoints was conducted. Patient demographics, reported symptoms at enrollment, and time between positive PCR test and start of intervention are shown in Table 1.

**Table 1:** Patient demographics and baseline characteristics by treatment group. Due to missing values of 4 participants that did not complete or fill in the intake survey, this table is based on data from 60 participants.

|  | Verum (n = 33 ) | Placebo (n = 27) |
| --- | --- | --- |
| Age (years) [mean, stdv] | 42 ± 12 | 43 ± 12 |
| Sex (female) [n, %] | 21 [62%] | 19 [63%] |
| Body Mass Index (BMI, kg/m<sup>2</sup>) [mean, stdv] | 26.6 ± 4.8 | 26.1 ± 5.5 |
| • Of which obesity (BMI >30) | 4 [12%] | 5 [19%] |
| Smoker (yes) [n, %] | 4 [12%] | 5 [19%] |
| Employment in |  |  |
| • Patient care [n, %] | 1 [3%] | 1 [4%] |
| • Children care [n, %] | 2 [6%] | 1 [4%] |
| • Teaching [n, %] | 7 [21%] | 5 [19%] |
| Inhalation allergy [n, %] | 15 [45%] | 7 [26%] |
| Lung disease (asthma, COPD) | 3 [9%] | 4 [15%] |
| Cardiac disease (yes) [n, %] | 2 [6%] | 0 [0%] |
| Immune disorder (yes) [n, %] | 1 [3%] | 1 [4%] |
| Diabetes (yes) [n, %] | 1 [3%] | 1 [4%] |
| Hypertension [n, %] | 3 [9%] | 4 [15%] |
| Days from positive PCR test [median, range] | 1 [-1,3] | 1 [0-4] |
| Days since onset of symptoms [median, range] | 3 [1-18] | 2 [1-11] |
| Fever (yes) [n, %] | 5 [15%] | 7 [26%] |
| Cough (yes) [n, %] | 24 [73%] | 17 [63%] |
| Sore throat (yes) [n, %] | 13 [39%] | 9 [33%] |
| Runny/blocked nose (yes) [n, %] | 20 [61%] | 22 [81%] |
| Shortness of breath (yes) [n, %] | 7 [21%] | 7 [26%] |
| Headache (yes) [n, %] | 21 [64%] | 18 [67%] |
| Loss of smell and taste (yes) [n, %] | 8 [24%] | 5 [19%] |
| Muscle pain (yes) [n, %] | 17 [52%] | 15 [56%] |
| Chills (yes) [n, %] | 13 [39%] | 7 [26%] |
| Fatigue (yes) [n, %] | 25 [76%] | 20 [74%] |
| Diarrhea (yes) [n, %] | 2 [6%] | 2 [7%] |

**Figure 1:**
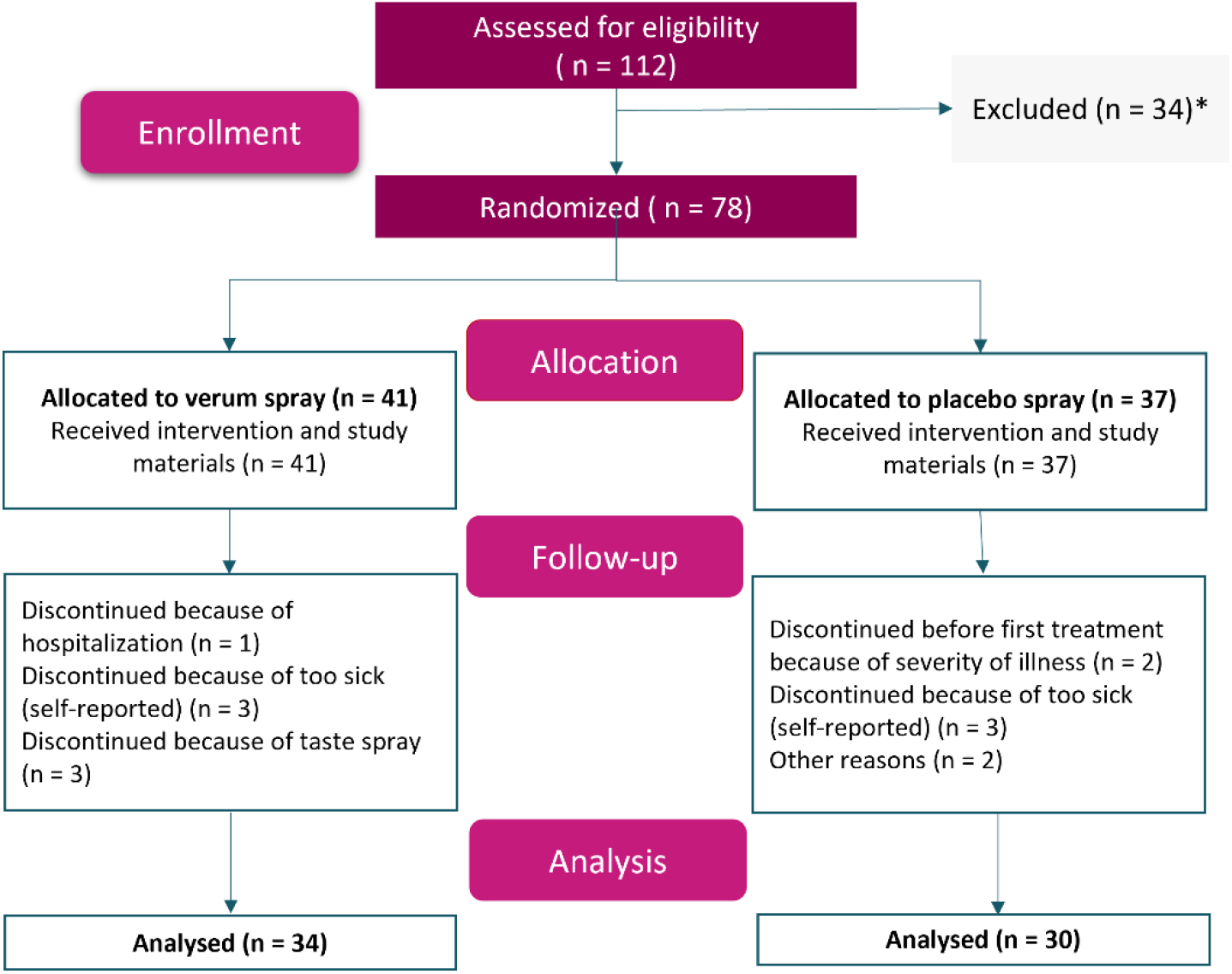
CONSORT flowchart for patient recruitment and enrollment. * Excluded for not meeting the inclusion criteria (mostly because time from positive PCR test was > 96 hours, often when people contacted the study coordinator in response to the press release message) or declined to participate after first contact with study coordinator. Of note, for the analysis of SARS-CoV-2 viral loads with RT-qPCR, 4 participants in the verum group and 3 participants in placebo already had a negative PCR test at T1 of our study. Hence, for this analysis, 30 participants and 27 participants were analyzed for verum and placebo, respectively.

The sprays were overall well-tolerated, but participants in both study groups often reported unpleasant taste (mainly verum group) or texture (verum and placebo) of the spray. Also for the online diaries, the compliance was high: the median number of completed diaries was 20/21 days, with 31.3% of the study population showing full compliance, with filling in the diary every days. The compliance of the self-sampling was 80.5% (509/632) for the combined nose/throat swabs, and 83.5% (132/158) for the fingerprick blood samples.

### Monitoring of symptoms in primary- care patients and impact of the microbiome-throat spray on symptom severity and time to improvement

Symptoms at the start of the study are shown in Table 1. Cough (68%), runny/blocked nose (70%), headache (65%) and fatigue (75%) were most frequently reported. The average total symptom score at start of the study was 13.4 ± 8.6 in the verum group and 15.2 ± 9.3 in the placebo group (difference not significant) (Table S1).

Severity of the symptoms was evaluated between both treatment groups over the study via the distribution of the different severity scores (total, URT, acute and symptom) at every day (see also Table S1). The same tendency for the verum and placebo group was observed with no significant differences (Figure 2A-D). Independent of treatment, raw symptoms scores showed high inter- and intra-individual fluctuation patterns (Figure S2), and scores were therefore propagated, smoothened and standardized (Figure S3). Furthermore, time to improvement was not significantly different between treatment groups and log odds (cox regression) were 0.125 ± 0.3, -0.003 ± 0.3, 0.111 ± 0.3, 0.58 ± 0.3 for total, system, URT and acute scores, respectively (Figure 2E-H). Over the entire study population, 59% of the individuals (independent of treatment) still experienced symptoms after 21 days. At this three-week timepoint, 5% reported acute symptoms, 39% systemic symptoms and 41% URT symptoms.

**Figure 2:**
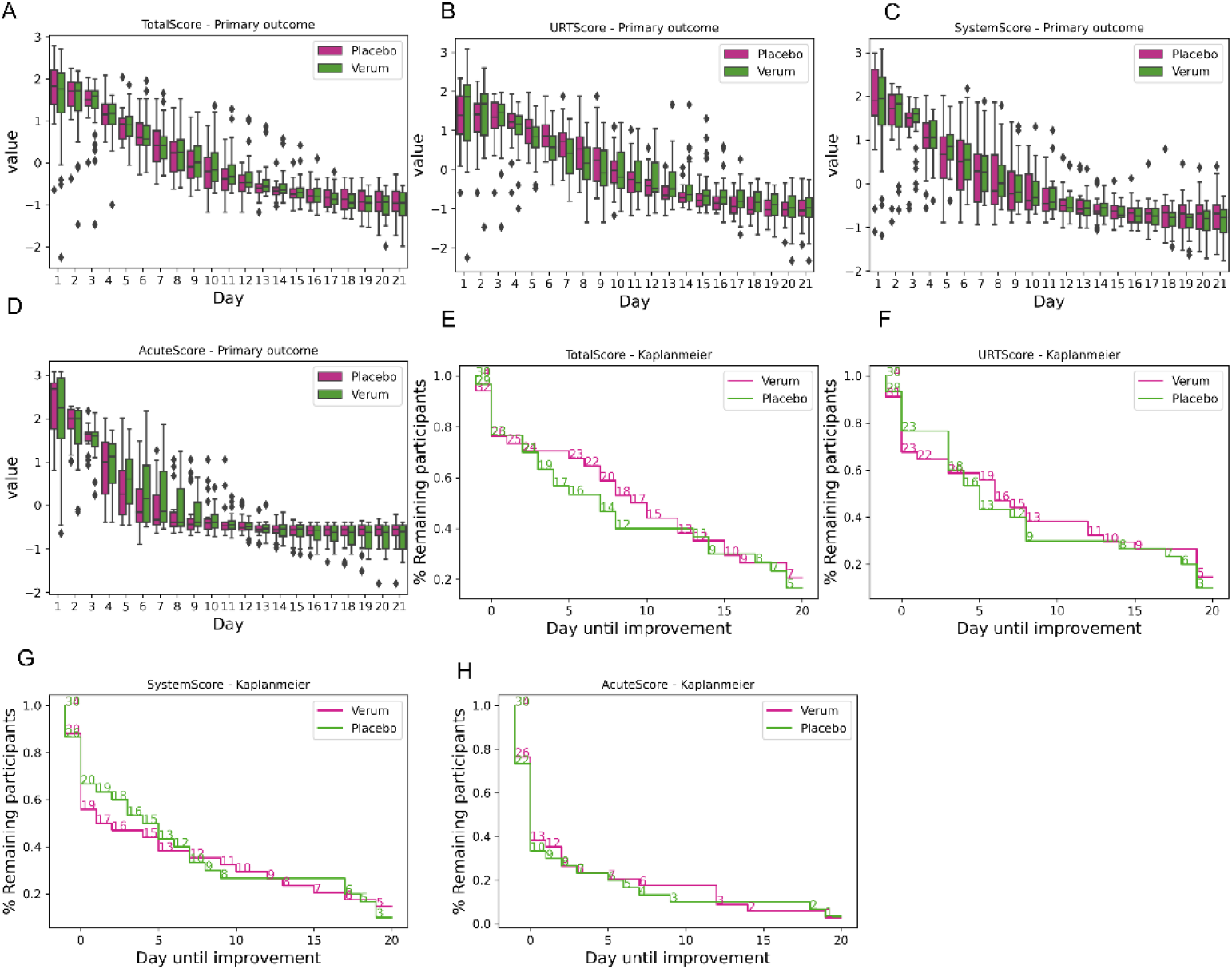
Symptom severity and time to improvement. Severity of the reported symptoms (A-D) was evaluated based on different scoring systems: Total score (A), URT score (B), system score (C) and acute score (D). Results are shown as standardized scores (z-score) to adjust for the highly subjective self-evaluation. Time to improvement (E-H) was also evaluated for the 4 scoring systems between both study groups. Survival analysis showed no significant differences for all tested scores between placebo and verum (p > 0.1).

### Impact of the microbiome spray on SARS-CoV-2 viral loads and relation with symptoms

At the start of the trial, 4/34 participants in the verum group and 3/30 participants in the placebo group had a negative RT-qPCR result despite testing positive less than 96h earlier. After one week, 73% of the participants in the verum group and 77% in placebo group tested positive (p = 1, Fisher’s exact test), while after 2 weeks this was 17% and 32%, respectively (p = 0.22, Fisher’s exact test). At the end of the trial, 2/30 (6.7%) patients in the verum group and 7/27 (26%) patients in the placebo group still tested positive (p = 0.07, Fisher’s exact test) (Figure 3A, 3B). Independent of the intervention, all symptoms had a strong correlation with being SARS-CoV-2 positive (Table S2), although several symptoms, such as cough, nasal symptoms and fatigue, were still reported upon a negative PCR result (Figure 3C).

**Figure 3:**
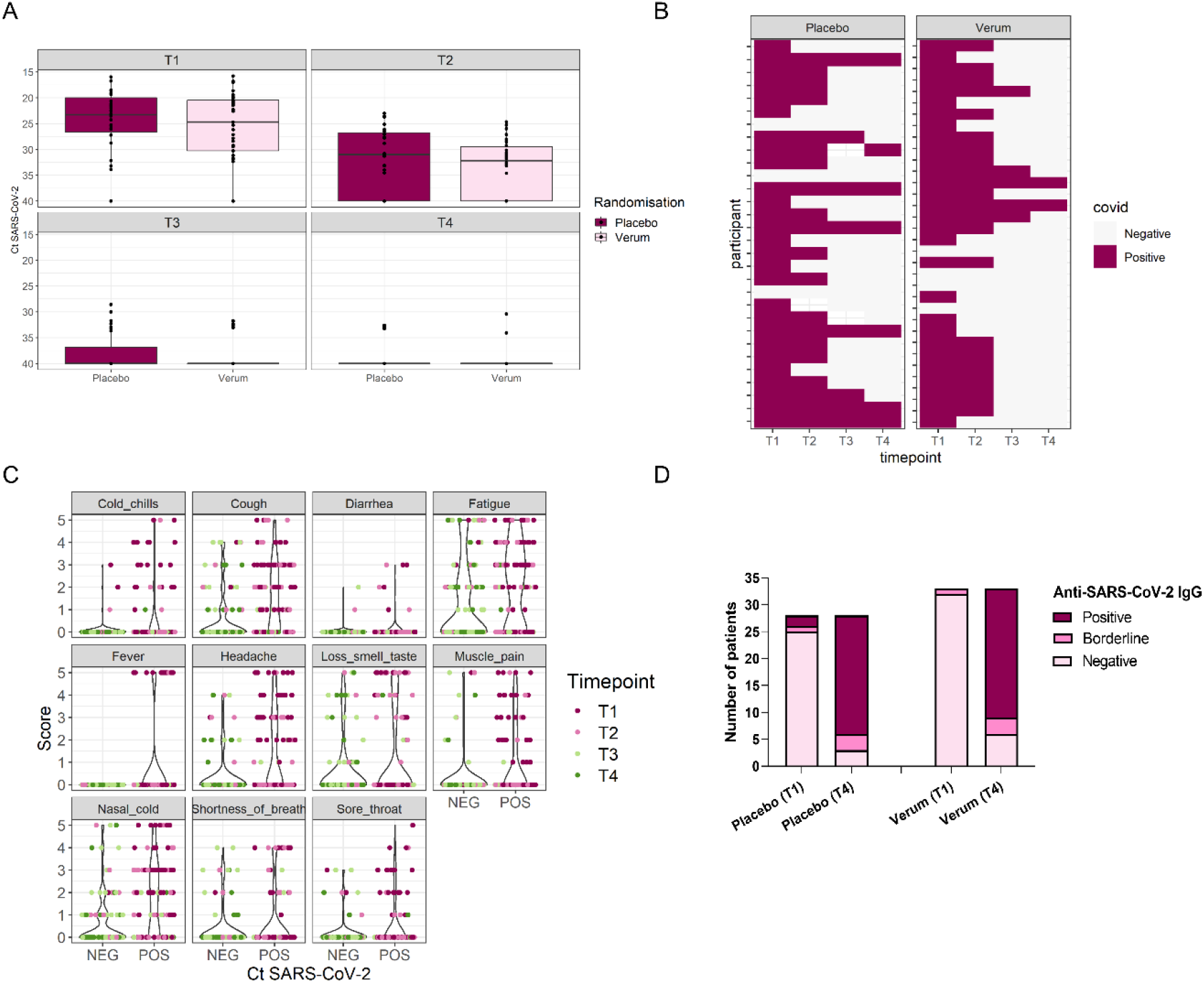
Viral loads in combined nose/throat swabs. A) SARS-CoV-2 viral loads in combined nose-throat swabs determined via PCR at start (T1), after 1 week (T2), after 2 weeks (T3) and after 3 weeks/end of the study (T4). Results are shown as Ct values. B) Heatmap showing presence or absence of SARS-CoV-2 based on positive PCR test. Each vertical line represents one participant. C) Relation between self-reported symptoms and SARS-CoV-2 viral load. D) Presence of anti-SARS-CoV-2 IgG antibodies in the blood of COVID-19 patients (COV) at the start (T1) and at the end (T4) of study comprising 3 weeks in between. Data for treatment groups verum and placebo is depicted as number of participants positive, borderline or negative for anti-SARS-CoV-2 IgG as part of the total number of participants per treatment group per time point. Only participants with blood samples available at both T1 and T4 were included in this analysis.

Analysis of self-collected fingerprick blood samples showed that at the start of the study, only 4/61 enrolled COVID-19 patients were positive or borderline positive for anti-SARS-CoV-2 IgG based on antibody reactivity against the receptor-binding domain (RBD), nucleocapsid (NCP) and spike proteins (S1S2) of SARS-CoV-2 [18] (Figure 3D). After 3 weeks, 51/61 patients were positive or borderline positive for anti-SARS-CoV-2 IgG, without significant differences between placebo and verum groups (p = 0.71, Chi-square test).

### Impact of SARS-CoV-2 infection on the upper airway microbiome

Principal coordinates analysis (PCoA) showed no major shifts in the overall nose/throat microbial composition for the time points nor for the microbiome treatment (Figure 4A). However, specific effects on abundances of certain taxa were observed. When focusing on abundances of the amplicon sequence variants (ASVs) of the *Lactobacillaceae* strains administered with the throat spray, significant differences were observed between verum and placebo groups at different time points with mean relative abundances for the *L. casei* ASV, *L. plantarum* ASV and *L. rhamnosus* ASV in the verum group found to be 1.6%, 1.3% and 0.5%, respectively, over the entire study (Figure 4B, Table S3). In the placebo group, these numbers were below 0.01% for all three ASVs (see also Table S3). Prevalences (presence) based on MiSeq data were 38.6%, 28% and 13.4% for the *L. casei* ASV, *L. plantarum* ASV and *L. rhamnosus* ASV, respectively, while this was 10.5%, 7% and 2% in the placebo group, pointing at the fact that the related taxa to the applied lactobacilli are also endogenously present, but in low numbers. Therefore, the presence of the specifically applied *Lactobacillaceae* strains was also confirmed via qPCR with clear difference between verum (on average detected in 82% of the study population) and placebo (on average 21%) and also between the estimated CFU/ml counts of the three strains, in a range of 10^8^ CFU/ml for *L. casei* AMBR2, 10^7^ CFU/ml for *L. plantarum* WCFS1, and 10^6^ CFU/ml for *L. rhamnosus* GG, in line with the concentrations in which they were added in the throat spray (Figure 4C, Table S4).

**Figure 4:**
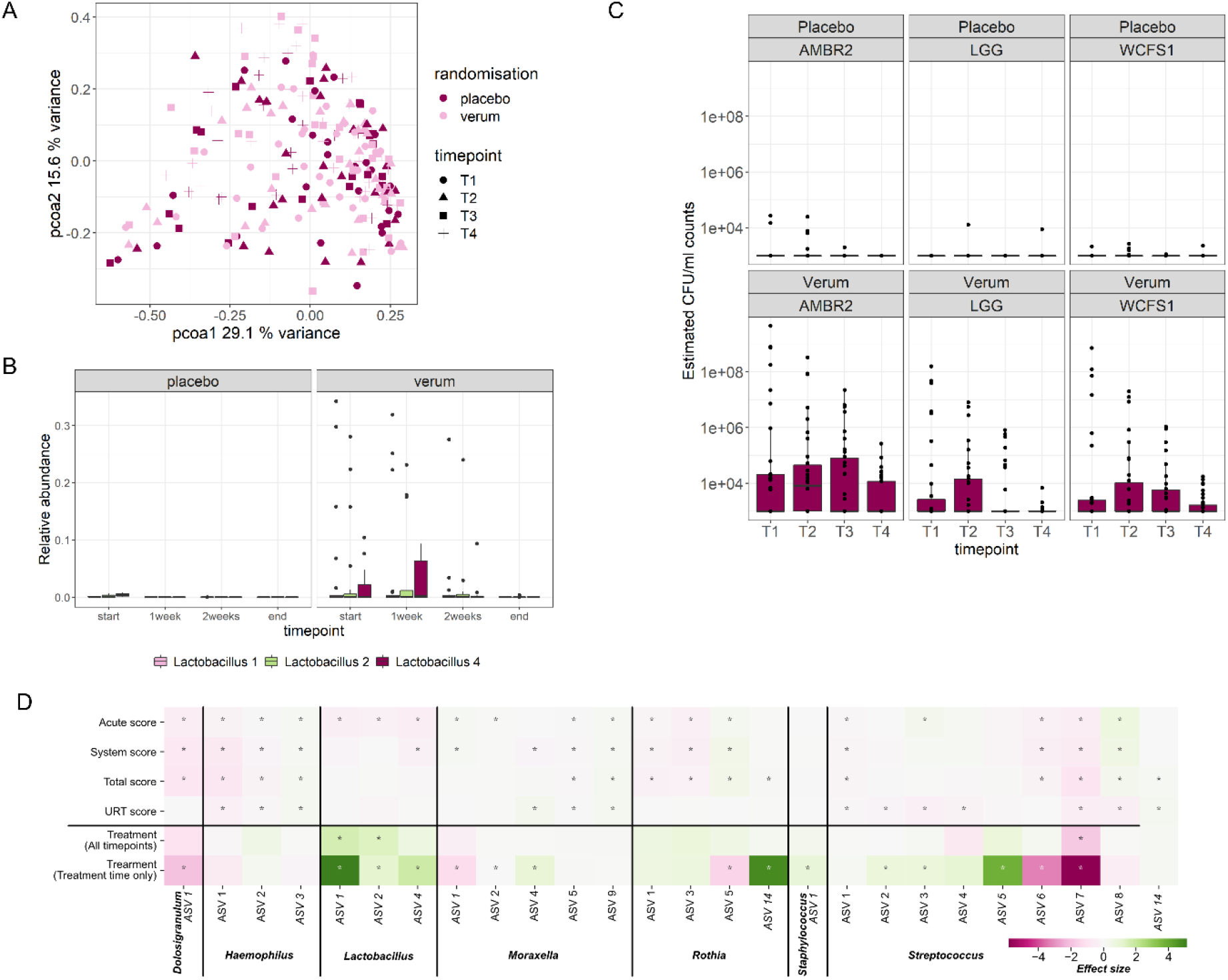
Microbial community composition in the airways (A), detection of the administered *Lactobacillaceae* strains (B-C) and correlation of treatment and symptom scores with bacterial taxa (D). A) PCoA was used to visualize the microbiome composition in combined nose/throat swabs for each treatment group and at the different timepoints. T1 = start, T2 = after 1 week, T3 = after 2 weeks, and T4 = end trial. B) Relative abundances of *L. casei* ASV, *L. plantarum* ASV and *L. rhamnosus* ASV between placebo and verum. See also Table S2 for mean relative abundances for all ASVs at the different timepoints and statistics. C) qPCR with species-specific primers for *L. rhamnosus* GG, *L. casei* AMBR2 and *L. plantarum* WCFS1 was used to estimate the CFU/ml counts. Based on the standard curve, the detection limit was estimated to be at 10^3^ CFU/ml. D) Correlation of treatment and symptom scores with bacterial taxa across all timepoints in the study. For the treatment, the correlation was also evaluated for treatment time only.

Next, we correlated the relative abundances of a selection of ASVs belonging to important airway genera (*Rothia, Dolosigranulum, Streptococcus, Staphylococcus, Haemophilus, Moraxella*, and *Lactobacillus*) with treatment, severity scores and viral load across all timepoints and made a further selection based on the ASVs that showed the highest effect sizes (Figure 4D). *Lactobacillus* ASV 1 (potentially *L. casei/paracasei/zeae*), *Lactobacillus* 2 (potentially *L. fabifermentans/paraplantarum/pentosus/plantarum*), and *Lactobacillus* ASV 4 (potentially *L. rhamnosus*) have a strong significant enrichment in the verum group, which was true across all timepoints and more pronounced on treatment timepoints only (Figure 4D). In addition to the deliberately added bacteria, other ASVs were positively correlated with the verum group, including *Moraxella* ASV 4 (*M. lacunata*), *Rothia* ASV 14 (*R. amarae*) and several commensal *Streptococcus* ASVs (among others *thermophilus, rubneri* and *sanguinis*). On the other hand, negative correlations with treatment were observed with the strongest effect sizes for *Dolosigranulum* ASV 1 (*D. pigrum*), *Streptococcus* ASV 7 (*S. gordonii*) and *Streptococcus* ASV 6 (*S. crispatus/oligofermentans/sinensis*).

Finally, positive and negative correlations between the symptom scores and specific taxa, with moderate effect sizes were found (Figure 4D). Of interest, a significant negative correlation was found for the ASVs corresponding to the applied lactobacilli and the acute symptom score, indicating that application of these lactobacilli could result in less acute symptoms. *Dolosigranulum* ASV 1, another lactic acid bacterium, had a negative correlation with the acute symptom score severity and even with the total score. Conversely, *Haemophilus* ASV 3 (*H. aegyptius*) positively correlated with the different symptom scores. Viral load did not have significant associations with specific taxa.

## Discussion

We evaluated the use of a specifically formulated topical microbiome throat spray with three selected members of the *Lactobacillaceae* in a placebo-controlled, remote self-sampling study in COVID-19 outpatients. Detailed microbiome and qPCR analysis showed a detection of the applied strains in the verum group in on average 82% of the participants based on qPCR with estimated concentrations between 10^6^-10^8^ CFU/ml. Analysis of the self-reported symptoms showed a patient-dependent disease progression with high intra- and interindividual variations and no significant effects of the intervention on the primary outcome in this rather small study population. However, a trend of faster decreasing viral loads was observed in the verum group compared to placebo, with after three weeks 6.7% and 26% of participants remaining positive based on RT-qPCR testing, respectively (p = 0.07). This remains to be substantiated in follow-up studies.

A first key finding of the study was the personal fluctuations of COVID symptoms, which we could document quite detailed thanks to online questionnaires that were developed within the Rapid European COVID-19 Emergency Response research (RECOVER) project [17]. Another study with daily monitoring of symptoms for two weeks also recently reported that the natural course of COVID-19 disease is highly patient-dependent and variable [19]. This highly fluctuating disease pattern and subjective self-evaluation of the symptom scores also complicates studies on treatments for COVID-19 symptoms. For instance, no effects of oral azithromycin in outpatients on the absence of self-reported symptoms after 14 days as primary outcome were observed in a study with 263 patients [20]. This was confirmed in the large open-label, multi-arm PRINCIPLE trial for the azithromycin group (n = 2265) [21]. On the other hand, inhaled budesonide showed positive effects in COVID-19 outpatients at risk aged 50-65 years (n = 4700) with a benefit in time to self-reported recovery of 2.9 days for patients in the budesonide group compared to usual care group [22]. While probiotic trials in COVID-19 outpatients are scarce compared to drug interventions, at least two recent studies reported symptom improvement. After intranasal administration of *Lactococcus lactis* W136 (n = 23), the proportion of patients with fatigue was lower in the verum group than placebo on day 7 (p = 0.02). In addition, patients in the verum group had reduced loss of sense of smell on day 9 (p = 0.03) and reduced shortness of breath on day 8 (p = 0.02) and day 12 (p = 0.04) compared to placebo [23]. However, it should be noted that the authors did not report any correction for multiple testing in this study, which is also not yet peer reviewed. In another larger trial (n = 300) with an oral probiotic mixture of *L. plantarum* strains and *Pediococcus acidilactici* KABP021, patients in the verum group reported fewer days of fever, cough, headache, body aches and shortness of breath [8].

Our results on the viral loads at day 21 are promising and in line with our previously reported *in vitro* data, where we showed that the microbiome members that were selected for the throat spray show clear antiviral effects against coronaviruses under controlled *in vitro* study set-ups [16]. In the trial with the oral probiotic *L. plantarum* strains and *Pediococcus acidilactici* KABP021, a significantly larger reduction in nasopharyngeal viral load was observed in the verum group at day 15 and day 30 (p < 0.001) [8]. Our data on viral loads should be further substantiated, because our study had several limitations. In addition to the already discussed high biological variation of COVID-19 disease progression and the rather small study population, we also experienced a suboptimal timepoint of intervention. The start point appeared too late after disease onset. This has also been observed in other trials with antivirals for respiratory infections [24,25]. For local applications of probiotics or microbiome therapeutics [6] compared to oral applications such as [8], it is especially important that the antiviral bacteria are provided early enough in the viral infection process, considering the more local mode of action. Oral probiotics target the gut and systemic immunity to reduce symptoms in later phases [26]. Here, we had to rely on the standard government PCR testing procedures in Belgium, resulting in a delay between testing and inclusion. Most participants started within 2-5 days after their first symptoms, when the viral loads were highest. A study design with a more preventive set-up might be more suitable to evaluate a microbiome throat spray with a mode of action early in the viral infection process. Moreover, we also observed that most patients still experienced symptoms at the end of the monitoring (5% still reported acute symptoms, 39% systemic symptoms and 41% URT symptoms), so that follow-up work with any potential therapeutic or nutritional interventions in outpatients preferably extents the follow-up period for the patients to evaluate long COVID effects.

Our finding of reduced viral loads also suggests some to-be-validated potential to reduce transmission to household members and other high-risk contacts. In a prospective cohort study conducted in the Netherlands and Belgium, it was shown that secondary transmission within households occurred in 44.4% of the households, mostly very early after the index patient was positive [17]. Moreover, it might be useful for future studies to stratify potential responders and non-responders based on the microbiome (e.g. exclude patients with high relative abundances of lactic acid bacteria *Dolosigranulum* because of the negative correlation found here with treatment), which is currently not standard in clinical trials with microbiome therapeutics or probiotics. Moreover, an alternative formulation such as a nose spray instead of the used throat spray, might be more favorable, since SARS-CoV-2 receptors are highly expressed in nasal epithelium, and the alpha variant that was dominant during the study has been shown to primarily target the nose [27]. In addition, based on in-house previous research, the selected lactobacilli have several beneficial modes of action such as antimicrobial and immunomodulatory properties, as well as barrier enhancing effects in the nasal epithelium [28–30]. However, this might also depend on the virus variant that is most dominant, because the current SARS-CoV-2 Omicron (B.1.1.529) variant seems to target the throat as the first or main site with high viral loads.

Since our study was one of the few in mild-to-moderate COVID patients outside the hospital and mostly based on self-sampling, we also generated relevant information irrespective of the treatment evaluated. In addition to the already mentioned intrapersonal fluctuations and interpersonal variations of disease symptoms, we also observed a robust detection of SARS-CoV-2 virus in self-collected combined nose/throat swabs. This exemplifies that a self-sampling approach for these types of samples is feasible, in line with the previously demonstrated effectivity of SARS-CoV-2 detection in oropharyngeal swabs collected by self-sampling during early infection to other read-outs and sample types [31]. Furthermore, our results indicate robust detection of IgG against the SARS-CoV-2 RBD, NCP and S1S2 antigens in self-collected dry blood spot samples, allowing detection of positive cases without the need for blood collection by healthcare professionals. Remote study set-ups with self-sampling and online questionnaires can thus represent a promising set-up for studies in COVID-19 outpatients and other infectious diseases. Collecting samples without involvement of a third party could reduce exposure, expand testing capacities, and minimize the burden on the hospitals and general practitioners during the COVID-19 pandemic [31,32].

Taken together, our study suggests that the microbiome throat spray might have beneficial effects via lowering nose/throat viral loads, potentially resulting in less virus transmission. Future studies are required to investigate this transmission to for instance household members. We also believe that a more preventive set-up is advised for the evaluation of this microbiome therapy, for instance for high-risk contacts.

## Supporting information

Figure 1: CONSORT flowchart for patient recruitment and enrollment

## Data Availability

All data produced in the present study are available upon reasonable request to the authors or will be deposited online after publication

## Acknowledgements

We would like to thank Sam Van Goethem for his advice on the study packages and logistics for the home sampling. We also would like to thank all GPs that helped with the recruitment and all study participants that participated in this trial. Furthermore, we want to thank the LebeerLab team, in particular Ines Tuyaerts, Nele Van de Vliet, Leen Van Ham, Marianne van den Broek, Sarah Ahannach, Vincent Greffe, and Marie Legein. Finally, we would like to thank Biobank Antwerpen (Antwerp, Belgium; ID: BE 71030031000).

