## Supplementary methods, Figure S1, Figure S2, Figure S3, Table S1, Table S2, Table S3, Table S4 for "Evaluation of a throat spray with lactobacilli in COVID-19 outpatients in a randomized, double-blind, placebo-controlled trial for symptom and viral load reduction"

**Participants:** COVID-19 outpatients, aged 18-65 years and that were not yet vaccinated against COVID-19, were recruited via general practitioners, local triage centers, and through a press release announcing the initiation of this study via newspaper and radio. Patients were contacted by the study coordinator within 96 hours of a positive PCR test to evaluate whether all inclusion and exclusion criteria were met. Upon inclusion, a study package with all materials required for the study was delivered at their place of self-isolation. The trial was conducted from February 24, 2021 to April 30, 2021 at the University of Antwerp. During this period, there were 241, 629 new cases confirmed in Belgium, which were dominated by the SARS-CoV-2 alpha-variant (between 58-86%) (Sciensano).

**Intervention**: Verum and placebo sprays were supplied by Yun NV (Niel, Belgium), and were indistinguishable in shape and color. The verum spray consisted of freeze-dried *Lacticaseibacillus casei* AMBR2, *Lactiplantibacillus plantarum* WCFS1, and *Lacticaseibacillus rhamnosus* GG in a ratio of 50%, 33.3%, and 16.7%, respectively, in sunflower oil with Aerosil, vitamin D3 and vitamin E. Placebo sprays had the same composition without the bacteria. Both verum and placebo sprays could be stored at room temperature. Quality control of the sprays and evaluation of the stability of the strains was performed at Yun NV and included microbial analyses, and evaluation of spray characteristics, sedimentation, viscosity and gelling at different temperatures from 4 to 25°C. Absence of non-lactobacilli was evaluated on CASO / sabouraud agar.

**Randomization and masking:** Randomization occurred in blocks of six patients with stratification for age and gender, using a randomization list generated with the Sealed Envelope web service (https://www.sealedenvelope.com/), by the responsible clinician, who was not involved in the outcome assessment and data analysis. All other researchers and doctors involved in the study were blinded. Participants were enrolled and assigned to study groups by the study coordinator at the time of the first contact via phone. If several household members participated in the study, randomization was done per household with treatment allocated based on the contact person of the household.

Verum and placebo products were given the label A or B provided together with the study package. Participants, investigators and outcome assessors were blinded for the treatment allocation until all analysis were performed and the study groups were unblinded by the formulation team.

**Symptom score evaluation and assessment of study compliance:** Ten common COVID-19 symptoms were monitored, including cough, sore throat, runny/blocked nose, shortness of breath, headache, muscle pain, chills, fatigue, loss of smell and taste, and fever. Each symptom was scored between 0 (no symptoms) and 5 (severe symptoms), according to [1], with the exception of fever, which received a binary score. Different symptom summary scores were compared: total score (sum of all reported symptoms), local URT score (the sum of cough, sore throat, and nasal discomfort scores), acute score (the sum of fever, diarrhea, chills, and muscle pain scores), and system score (the sum of fever, shortness of breath, muscle pain, chills, fatigue, and diarrhea scores). The symptomatic tipping point was used to determine time to improvement. Therefore, the time interval after which a particular symptom only improved was determined for each participant, *i.e*. after that time interval, the symptom score was continuously decreasing.

Study compliance was assessed based on the responses via the online diary, the self-sampling of combined nose throat swabs and blood fingerprick samples, and the use of the spray. The latter was evaluated via self-reporting and the spray bottles were weighted by the study team.

**Sample collection**: Samples were collected via self-sampling with guided instruction books and videos. Combined nose/throat swabs for microbiome analysis and determination of SARS-CoV-2 viral loads were collected every week from start for a total of 4 times. Blood fingerprick samples (dried blood spots) to analyze SARS-CoV-2 IgG antibodies were self-sampled at the start and end of the trial. All samples were stored at -20°C for swabs or 4°C in case of the blood fingerpricks.

**Analysis of SARS-CoV-2 viral loads and SARS-CoV-2 IgG antibodies**: Combined nose/throat swabs were collected with eNAT™ swabs and stored at -20°C at the participant's house until all samples were collected at the end of the study (approximately 4 weeks after the start of the study). Samples for RNA extractions were transported to the Antwerp University Hospital (Clinical Biology, prof. H. Goossens). RNA extractions were done with the EASYMAG® kit with prior proteinase K treatment. The applied duplex real-time PCR is based on the publication of Corman and colleagues [2] and on the detection of *RNAseP* as described in the CDC-protocol "Real-time RT-PCR panel for detection of 2019-Novel Coronavirus, CDC, instructions for use". Both assays are combined in the real-time duplex assay running under the conditions of the former assay. For SARS-CoV-2 IgG antibodies analysis, blood fingerprick samples were stored on Whatmann filterpaper at -20°C or 4°C and analyzed via an in-house developed Luminex bead-based assay targeting antibodies against the receptor-binding domain (RBD), nucleocapsid (NC) and spike (S1S2) proteins of the SARS-CoV-2 Wuhan strain [3,4].

**Bacterial DNA extraction from combined nose/throat swabs and Illumina MiSeq 16S rRNA amplicon sequencing:** Self-collected combined nose/throat swabs were stored at -20°C at the participants' house until the end of the study. After transportation on ice, samples for DNA analysis were kept at -20°C in the lab until further processing. Prior to DNA extraction, all samples were vortexed 15-30 seconds and 500 µL of the eNAT buffer was used for automatic extraction using DNeasy 96 PowerSoil Pro QIAcube HT Kit (QIAGEN). Negative extraction controls were included at regular time points throughout the study. All samples were eluted with 100 µL elution buffer and DNA concentrations were measured using the Qubit 3.0 Fluorometer (Life Technologies, Ledeberg, Belgium).

Amplicon sequencing (V4 region of the 16S rRNA gene) was performed using an in-house optimized protocol [5,6]. Processing and quality control of the reads was performed using the R package DADA2, version 1.6.0. All data handling and visualization was performed in R version 3.4.4 using the tidyverse set of packages and the in-house package tidyamplicons, version 0.2.1 (publicly available at github.com/SWittouck/tidyamplicons), as described in [5].

**Detection of administered *Lactobacillaceae* strains using qPCR:** Specific primers for *L. casei* AMBR2, *L. plantarum* WCFS1 and *L. rhamnosus* GG were used for qPCR cfr. [7]. Four µL of each extracted DNA sample was combined with 10 µL Power SYBR Green PCR Master Mix, 0.3 µL of each primer (20 µM) and 5.4 µL of RNase-free water. The cycle threshold (Ct) value of each sample was used to calculate the concentration of the strain present in the sample. Non-template controls were included for each run.

**Sample size:** Studies that have tested the administration of beneficial lactobacilli into airways through a throat spray to exert respiratory effects and antiviral activity are very limited. Therefore, it was difficult to calculate the capacity for this type of potency studies. Therefore, we looked for studies using other probiotic formulations in patients with COVID-19. Based on the available literature at the time the study was designed, we aimed to recruit at least 150 individuals, 75 in each group. This sample size allows to demonstrate a difference in mean symptom score of 0.6 times the standard deviation (effect size 0.6) between the two study groups (level of significance=5%; power=80%) taking into account 10% dropout in each study group. Recruitment was however stopped earlier (n = 78) due to difficulties to find sufficient study participants due to a drop in infection cases and beginning of broad vaccination campaigns in Belgium.


[1] Verberk J, de Hoog M, Westerhof I, van Goethem S, Lammens C, Ieven M, et al. Transmission of SARS-CoV-2 within households: A prospective cohort study in the Netherlands and Belgium - Interim results. MedRxiv 2021:1–18. https://doi.org/10.1101/2021.04.23.21255846.

[2] Corman VM, Landt O, Kaiser M, Molenkamp R, Meijer A, Chu DKW, et al. Detection of 2019 novel coronavirus (2019-nCoV) by real-time RT-PCR. Eurosurveillance 2020;25. https://doi.org/10.2807/1560-7917.ES.2020.25.3.2000045.

[3] Mariën J, Ceulemans A, Bakokimi D, Lammens C, Ieven M, Heytens S, et al. Prospective SARS-CoV-2 cohort study among primary health care providers during the second COVID-19 wave in Flanders, Belgium. Fam Pract 2021. https://doi.org/10.1093/FAMPRA/CMAB094.

[4] Mariën J, Ceulemans A, Michiels J, Heyndrickx L, Kerkhof K, Foque N, et al. Evaluating SARS-CoV-2 spike and nucleocapsid proteins as targets for antibody detection in severe and mild COVID-19 cases using a Luminex bead-based assay. J Virol Methods 2021;288:114025.

https://doi.org/10.1016/J.JVIROMET.2020.114025.

[5] De Boeck I, Wittouck S, Wuyts S, Oerlemans EFM, van den Broek MFL, Vandenheuvel D, et al. Comparing the Healthy Nose and Nasopharynx Microbiota Reveals Continuity As Well As Niche-Specificity. Front Microbiol 2017;8:2372. https://doi.org/10.3389/fmicb.2017.02372.

[6] De Boeck I, Wittouck S, Martens K, Claes J, Jorissen M, Steelant B, et al. Anterior Nares Diversity and Pathobionts Represent Sinus Microbiome in Chronic Rhinosinusitis. MSphere 2019;4:e00532-19. https://doi.org/10.1128/mSphere.00532-19.

[7] Spacova I, De Boeck I, Cauwenberghs E, Delanghe L, Bron PA, Henkens T, et al. Live biotherapeutic throat spray for respiratory virus inhibition and interferon pathway induction. BioRxiv 2022:2022.01.25.477549. https://doi.org/10.1101/2022.01.25.477549.

### Supplementary Figures and tables

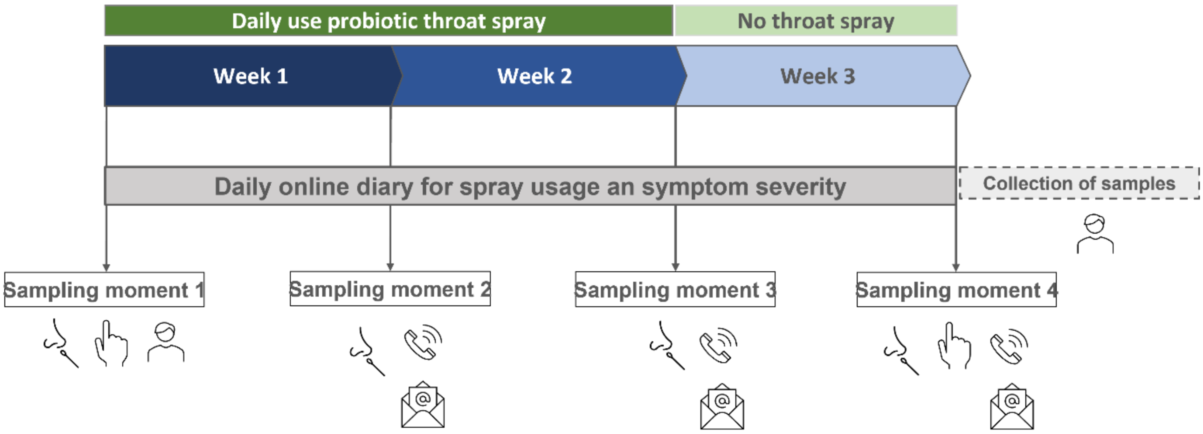


**Figure S1: Overview of the study set-up.** Patients were asked to use the verum spray or placebo for 14 days, with a recommended use of 5 times a day, which was also monitored via the online diary. After the intervention, patients were followed-up for an additional 7 days (so in total 21 days of symptom follow-up). They were asked to fill in an online diary via Qualtrics (Qualtrics, Provo, UT, USA) for each day of the study reporting spray usage as well as symptom severity. Every week, patients were contacted via teleconsults or email. At the end of the study, the study coordinator planned a final visit to collect all samples. Combined nose/throat swabs were collected at start (T0), after 1 week (T1), after 2 weeks (T2) and at the end of the trial (T3). Fingerprick blood samples were collected at T0 and T3.

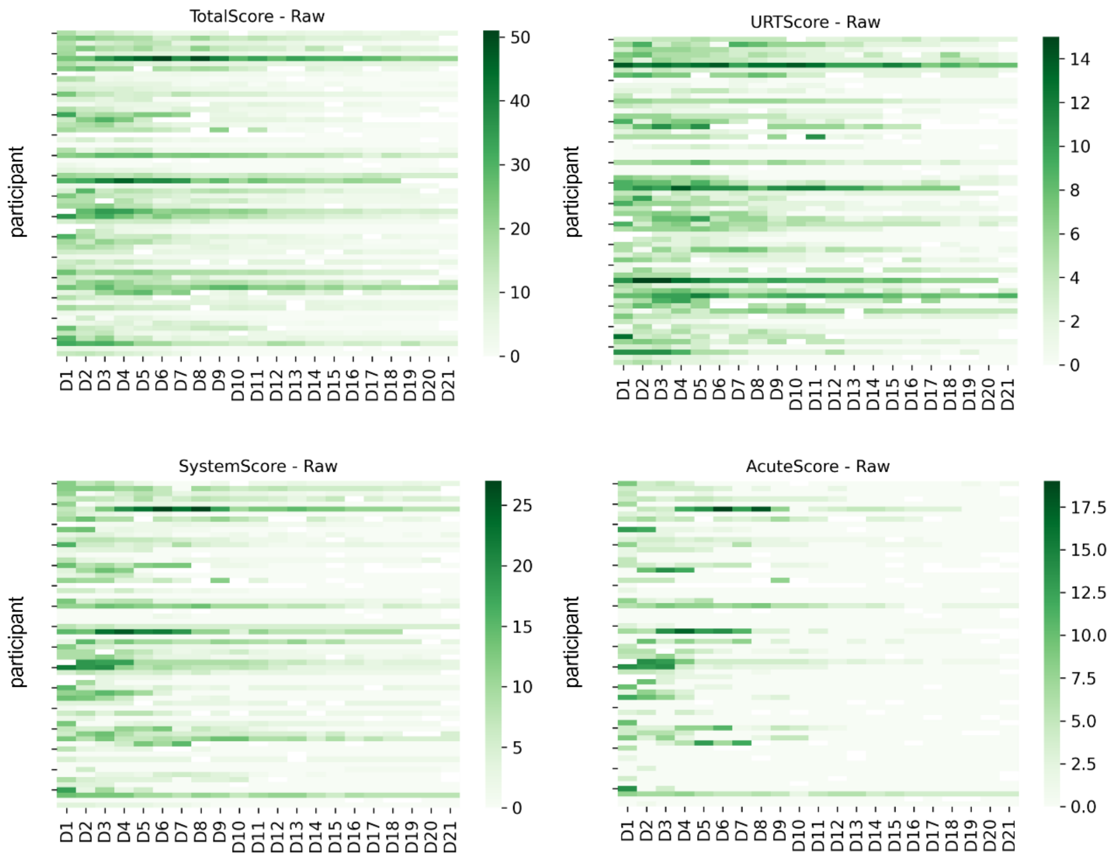


**Figure S2: Raw symptom scores for the different scoring systems.** Each vetrtical line represents one participant.

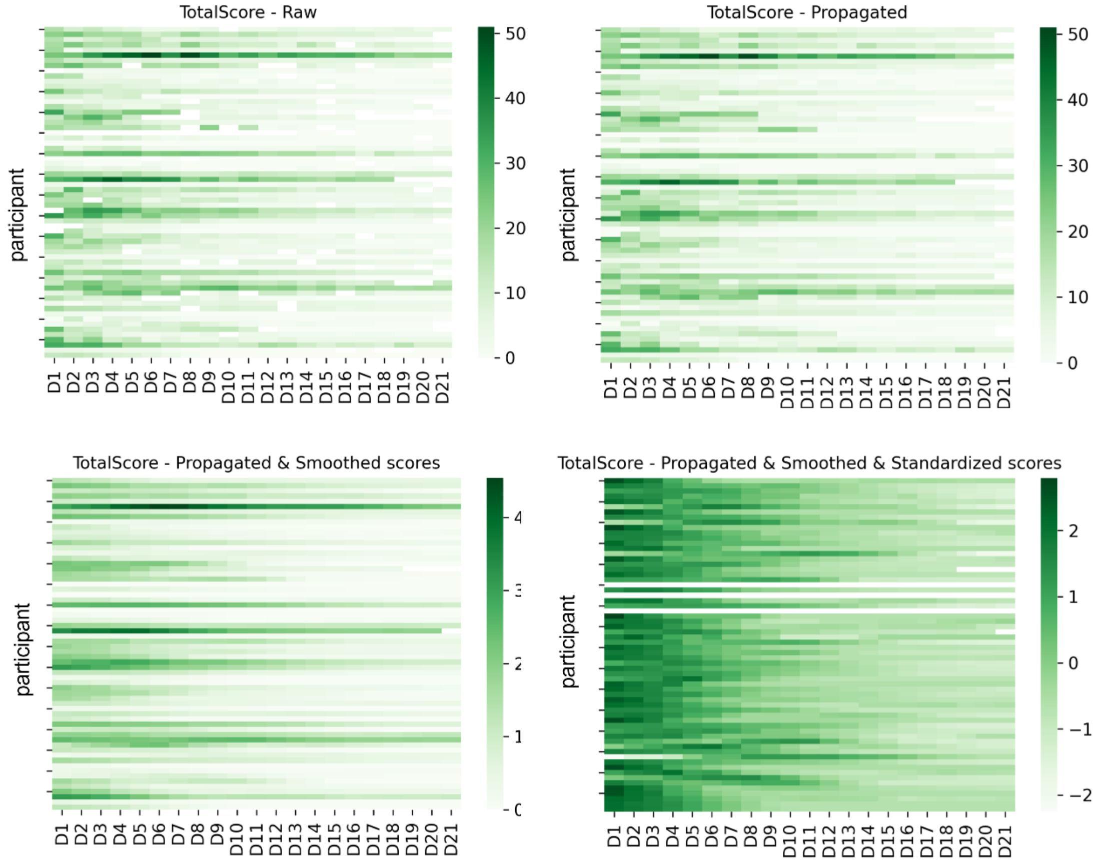


**Figure S3: Example of symptom processing for the total score**. Raw scores were first propagated, next smoothed and finally standardized (z-scores). Each vertical line represents one participant. The same analysis was done for the URT score, acute score and system score.

**Table S1: Spray usage and absolute severity score in both study groups.** Spray usage is depicted by the number of days the sprays were used (self-reported) and the remaining weight of the sprays (measured by the study team). Absolute severity scores (means ± stdev) for the different symptom summary scores are shown: total score (sum of all reported symptoms), local URT score (the sum of cough, sore throat, and nasal discomfort scores), acute score (the sum of fever, diarrhea, chills, and muscle pain scores), and system score (the sum of fever, shortness of breath, muscle pain, chills, fatigue, and diarrhea scores).

| | **Verum (n = 33 )** | **Placebo (n = 27)** |
|---|---|---|
| **Sprays used: number of days [mean, stdv]** | 19 ± 3 | 19 ± 3 |
| **Sprays used: remaining weight (grams)[mean, stdv]** | 72.2 ± 5.4 | 71.7 ± 6 |
| **Total score [mean, stdv]** | | |
| • T1 | 13.4 ± 8.6 | 15.2 ± 9.3 |
| • T2 | 10 ± 10.3 | 7.2 ± 6.9 |
| • T3 | 6 ± 7 | 3.1 ± 4 |
| • T4 | 4.3 ± 5.7 | 1.8 ± 3.1 |
| **URT score [mean, stdv]** | | |
| • T1 | 4.3 ± 3.4 | 5.3 ±3.2 |
| • T2 | 3.3 ± 3.2 | 3.1 ± 2.7 |
| • T3 | 2.3 ± 3 | 1.2 ± 2 |
| • T4 | 1.5 ± 2.5 | 0.39 ± 0.6 |
| **Acute score [mean, stdv]** | | |
| • T1 | 3 ± 3 | 3.3 ± 3.5 |
| • T2 | 1.4 ± 3.1 | 0.6 ± 1.3 |
| • T3 | 0.3 ± 0.8 | 0.2 ± 0.7 |
| • T4 | 0 ± 0 | 0.2 ± 0.5 |
| **System score [mean, stdv]** | | |
| • T1 | 6.1 ± 4.7 | 6.8 ± 5.4 |
| • T2 | 4.4 ± 5.4 | 2.7 ± 3.2 |
| • T3 | 2.3 ± 3.5 | 1.3 ± 1.9 |
| • T4 | 1.6 ± 2.2 | 1 ± 1.7 |

**Table S2: Correlation symptoms and being SARS-CoV-2 positive**. A random effect model was used (symptom ~ covid + (1|person)).

| index | Estimate | Std. Error | df | t value | Pr(>\|t\|) | test | qvalue |
|---|---|---|---|---|---|---|---|
| covidTrue | 0.314200 | 0.073315 | 1260.486581 | 4.285644 | 1.960797e-05 | Q3_2.Score_Keelpijn | 2.941196e-04 |
| covidTrue | 0.428104 | 0.075965 | 1268.742822 | 5.635510 | 2.148318e-08 | Q3_7.Score_Koude_rillingen | 3.222477e-07 |
| covidTrue | 0.476495 | 0.106387 | 1259.190851 | 4.478868 | 8.184613e-06 | Q3_3.Score_Neusverkouden | 1.227692e-04 |
| covidTrue | 0.477897 | 0.085126 | 1257.967353 | 5.613977 | 2.430525e-08 | Q3_1.Score_Hoesten | 3.645787e-07 |
| covidTrue | 0.494028 | 0.089678 | 1272.280454 | 5.508887 | 4.365740e-08 | Q2_1.Score_Koorts | 6.548609e-07 |
| covidTrue | 0.530083 | 0.102556 | 1260.710605 | 5.168725 | 2.738120e-07 | Q3_6.Score_Spierpijn | 4.107181e-06 |
| covidTrue | 0.578874 | 0.106427 | 1257.838613 | 5.439154 | 6.426810e-08 | Q3_8.Score_Vermoeidheid | 9.640215e-07 |
| covidTrue | 0.822822 | 0.107083 | 1261.759231 | 7.683934 | 3.083800e-14 | Q3_5.Score_Hoofdpijn | 4.625700e-13 |
| covidTrue | 1.266727 | 0.189839 | 1258.037498 | 6.672655 | 3.755642e-11 | URTScore | 5.633463e-10 |
| covidTrue | 1.494129 | 0.221934 | 1264.108524 | 6.732307 | 2.527026e-11 | AcuteScore | 3.790539e-10 |
| covidTrue | 2.237859 | 0.297700 | 1258.972580 | 7.517170 | 1.058896e-13 | SystemScore | 1.588344e-12 |
| covidTrue | 4.420475 | 0.511728 | 1258.079581 | 8.638323 | 1.702141e-17 | TotalScore | 2.553211e-16 |

**Table S3: Mean relative abundances of *Lactobacillaceae* ASVs** administered via the throat spray at the different timepoints in verum and placebo. A Wilcoxon test was used to evaluate statistical differences between verum and placebo group.

| ***Lactobacillaceae*** **ASV** | **Timepoint** | **Mean relative abundance in verum** | **Mean relative abundance in placebo** | **p-value** |
|---|---|---|---|---|
| *L. casei* ASV | T1 | 0.026905287 | 0.0001976367 | 0.30 |
| | T2 | 0.025694896 | 0.0001236477 | 0.056 |
| | T3 | 0.010757218 | 8.007239e-06 | 0.001 |
| | T4 | 0.0001428949 | 2.530286e-05 | 0.37 |
| *L. plantarum* ASV | T1 | 0.022253851 | 0.0002801401 | 0.53 |
| | T2 | 0.019241153 | 0.0001108163 | 0.12 |
| | T3 | 0.009505836 | 0.000000e+00 | 0.005 |
| | T4 | 0.0001831164 | 8.910997e-05 | 0.24 |
| *L. rhamnosus* ASV | T1 | 0.007690299 | 0.0003379468 | 1 |
| | T2 | 0.007265016 | 0.0000000000 | 0.18 |
| | T3 | 0.003396812 | 0.000000e+00 | 0.32 |
| | T4 | 0.0000000000 | 5.718128e-06 | 0.006 |

**Table S4: Average estimated CFU/ml counts for the applied *Lactobacillaceae*** in verum and placebo based on qPCR and % of participants were the strains were detected. Of note, also in the placebo group we observed some detection, but this is probably because these lactobacilli are also present as endogenous members in low numbers in the respiratory tract.

| **Strain** | **Estimated CFU/ml verum** | **Estimated CFU/ml placebo** | **% of participants detected verum** | **% of participants detected placebo** |
|---|---|---|---|---|
| *L. casei* AMBR2 | $1.1 \times 10^8$ | $9.1 \times 10^3$ | 85% (29/34) | 23% (7/30) |
| *L. rhamnosus* GG | $6 \times 10^6$ | $7.5 \times 10^3$ | 79% (27/34) | 10% (3/30) |
| *L. plantarum* WCFS1 | $1.7 \times 10^7$ | $1.5 \times 10^3$ | 82% (28/34) | 30% (9/30) |
